# A novel mutation in EROS (*CYBC1*) causes Chronic Granulomatous Disease

**DOI:** 10.1101/2023.04.21.23286144

**Authors:** Paige M Mortimer, Esme Nichols, Joe Thomas, Rachna Shanbhag, Neha Singh, Eve L Coomber, Talat H Malik, Matthew C Pickering, Lyra Randzavola, William Rae, Sagar Bhattad, David C Thomas

## Abstract

Chronic Granulomatous Disease (CGD) is an inborn error of immunity characterised by opportunistic infection and sterile granulomatous inflammation. CGD is caused by a failure of reactive oxygen species (ROS) production by the phagocyte NADPH oxidase. Mutations in the genes encoding phagocyte NADPH oxidase subunits cause CGD. We and others have described a novel form of CGD (CGD5) secondary to lack of EROS (*CYBC1*), a highly selective chaperone for gp91*phox*. EROS-deficient cells express minimal levels of gp91*phox* and its binding partner p22*phox*, but EROS also controls the expression of other proteins such as P2×7. The full nature of CGD5 is currently unknown. We describe a homozygous frameshift mutation in *CYBC1* leading to CGD. Individuals who are heterozygous for this mutation are found in South Asian populations (allele frequency = 0.00006545), thus it is not a private mutation. Therefore, it is likely to be the underlying cause of other cases of CGD.

## Introduction

The multi-subunit phagocyte NADPH oxidase generates reactive oxygen species (ROS) and is crucial for host defence ^1^. Deficiencies in individual subunits (gp91*phox*, p22*phox*, p47*phox*, p67*phox* and p40*phox*) of the phagocyte NADPH oxidase cause chronic granulomatous disease (CGD), a severe inborn error of immunity (IEI) characterised by severe infections with catalase positive organisms. Life threatening infections with *Staphylococcus, Salmonella* species, *Burkholdheria* and *nocardia* are all well documented. Fungal infections with species such as *Aspergillus* are also common and are associated with severe morbidity. While better anti-microbial prophylaxis and treatment accompanied by the advent of bone marrow transplantation for CGD has improved outcomes, it remains a severe disease that requires prompt identification and treatment. We previously showed that EROS/*CYBC1* is essential for the generation of ROS because it is the chaperone for the membrane-bound gp91*phox*-p22*phox* heterodimer in both mouse and humans ^2–4^. We and others have shown that mutations in EROS/*CYBC1* can cause CGD5 ^3,5^. The small number of cases described so far appear to have some typical features of CGD, such as opportunistic infections and inflammatory bowel disease, but also some features not usually associated with CGD including; autoimmune haemolytic anaemia, chronic glomerulonephritis and even some viral infections. Thus far, the only pathogenic EROS/*CYBC1* mutations identified are private mutations restricted to one family ^3^ (as a result of consanguineous marriage) or are limited to the Icelandic population ^5^. We describe a novel pathogenic mutation in EROS/CYBC1 that is likely to account for more cases worldwide as awareness of CGD5 grows.

## Results/Case History

A female in her twenties has been unwell for three years. Her childhood history is notable for tuberculosis in the first five years of life and an episode of uveitis during teenage years, which resolved following topical steroid treatment. She was otherwise well. At the onset of symptoms she developed a prolonged cough and fever. She was treated for radiographically proven pneumonia (**Figure 1A**) and responded to oral antibiotics. However, she developed a dry cough and shortness of breath 3 months later. Two weeks into the illness, she developed a rash over the trunk, dry mouth and swelling of her cervical lymph nodes. A biopsy of the skin rash showed erythema annulare centrifugum and fine needle aspiration cytology of the neck nodes showed granulomatous lesions. Testing by Mantoux and TB gold was negative. A CT scan of her chest showed upper zone nodules and ground glass opacities. ACE2 was elevated at 122 (12-68 units/ml). With a working diagnosis of sarcoidosis she was started on prednisolone (40 mg) and her respiratory symptoms, rash and neck nodes resolved. The steroids were gradually tapered, and methotrexate was added but then stopped due to liver dysfunction.

**Figure 1:**
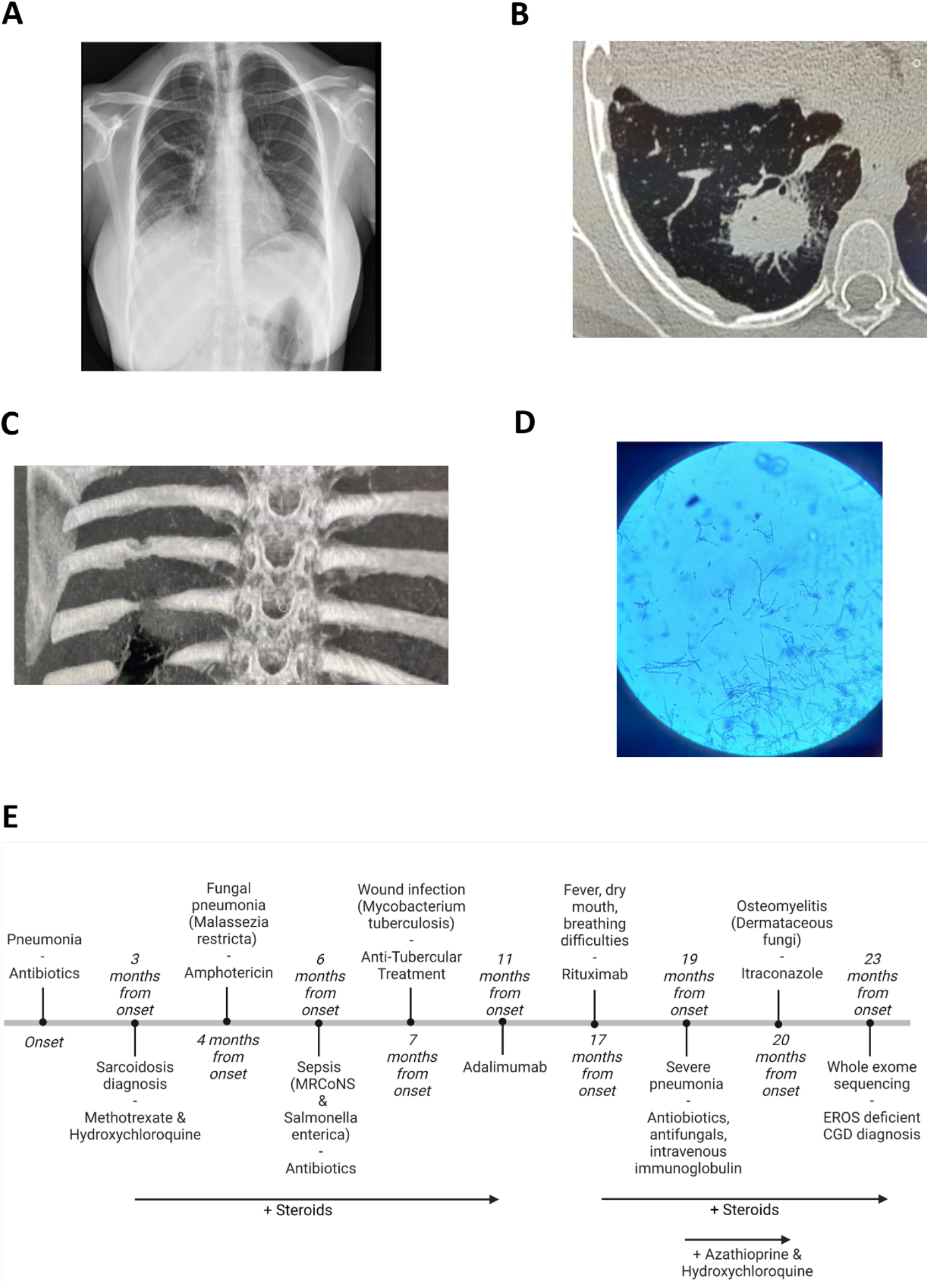
Clinical history of the patient. **(A)** Chest x-ray of pneumonia. **(B)** PET CT scan of a large necrotic lesion in the right lung with infiltration into the pleura. **(C)** CT scan of right 7th rib osteomyelitis near previous surgical wound. **(D)** Dematiaceous fungi growth from excised rib. **(E)** Timeline of the patient’s clinical course.

Between 4 and 7-months post-onset she was admitted to hospital three times for episodes of dry cough and shortness of breath. She developed fever and chest pain when steroids were reduced to 15 mg daily. A new CT chest showed traction bronchiectasis, multiple peri-lymphatic and parenchymal nodules and a minimal right pleural effusion. Bronchoalveolar lavage was negative for a TB panel, fungal stain and culture. Given the complexity of her presentation, a PET CT was undertaken. It showed a large necrotic lesion in the right lung with infiltration into the pleura (**Figure 1B**). A segmental resection of the involved lung by transthoracic surgery was carried out. This demonstrated dematiaceous fungi (Malasezzia restricta). She was treated with amphotericin B and steroids were continued. A blood culture at this time also demonstrated methicillin resistant coagulase negative staphylococci. During this period, she had ongoing liver dysfunction and a liver biopsy showed granulomatous hepatitis. She then developed fever and infection with *Salmonella* species was found on blood culture. A bone marrow culture also reported *Salmonella* species. A bone marrow biopsy showed granulomatous infiltration. In view of the granulomatous infiltration in multiple organs, anti-tubercular therapy (ATT) was started. She had ongoing fevers in the next month, despite four weeks treatment with amphotericin and hence, amphotericin was stopped. Her dose of steroid was increased to 1mg/kg prednisolone. With a combination of steroids and ATT, there was resolution of fever and cough.

At 12 months from onset, the surgical wound caused by the lung biopsy failed to heal and there was intermittent pus discharge from the wound. At this time, culture from the wound showed growth of Mycobacterium tuberculosis (45-day culture) and therapy was changed from modified ATT (in view of hepatitis and concurrent Voriconazole treatment) to standard ATT (6 months modified ATT followed by 9 months standard ATT), following which the wound healed. However, on reducing steroids there was recurrence of fever. Steroid sparing agents like mycophenolate mofetil and azathioprine were trialled, with no response. She also developed cushingoid features hence injected anti-TNF-α therapy (Adalimumab) was started 11 months post-onset, given for 6 months and stopped 16 months post-onset. Corticosteroids were tapered and stopped 15 months post-onset. She had a recurrence of fever at 17 months from onset and was treated with steroids and 2 doses of injected rituximab for a presumed flare of an autoinflammatory manifestation. At 19 months post-onset she developed fever and severe cough with hypoxia requiring ventilation. She was treated for atypical *stenotrophomonas maltophila* pneumonia with antibiotics, anti-fungals and intravenous immunoglobulin. She remained well for the next year on steroids, azathioprine and hydroxychloroquine. At 31 months post-onset she developed recurrence of fever and cough. A CT scan of the chest showed right 7th rib osteomyelitis near the previous surgical wound (**Figure 1C**). A rib excision was carried out that again showed dematiaceous fungi (Cladophialophora spp, **Figure 1D**) and she was treated with itraconazole for 1.5 months. A summary of her clinical course is shown in **Figure 1E**.

Given her unusual presentation with multiple episodes of severe infection and inflammation, she underwent whole exome sequencing. This demonstrated a homozygous mutation in the *CYBC1* gene (rs1291759701), Exon 7, c.327dup (p.Val110CysfsTer40), predicted to be pathogenic and a provisional diagnosis of diagnosis of CGD was established. This was confirmed by Sanger sequencing (**Figure 2A**). There were no deleterious mutations in known NADPH oxidase subunits. This mutation results in the insertion of a nucleotide that has two linked consequences:

**Figure 2:**
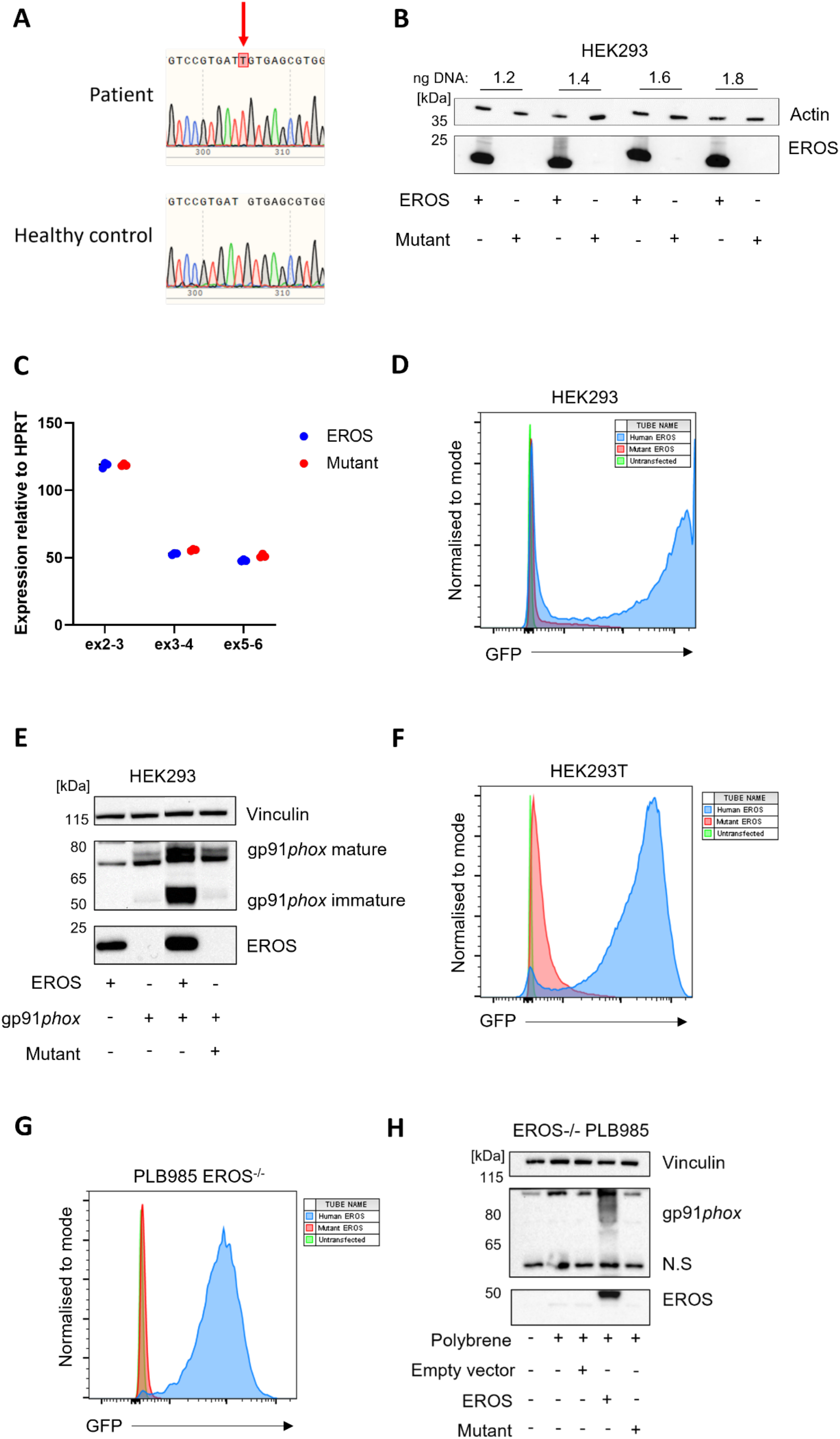
Novel EROS mutation abrogates EROS and cannot support gp91*phox* expression. **(A)** Sanger sequencing of CYBC1 in patient carrying the c.327dup (p.Val110CysfsTer40) mutation. **(B)** Normal and c.327dup (p.Val110CysfsTer40) EROS DNA was transfected into HEK293 cells and EROS protein expression measured by western blot and **(C)** gene expression measured by qPCR. EROS probes targeting several exon junctions were used. EROS is shown relative to HRPT. **(D)** GFP-tagged c.327dup (p.Val110CysfsTer40) and normal EROS DNA were transfected into HEK293 cells and analysed by flow cytometry. **(E)** Normal or c.327dup (p.Val110CysfsTer40) EROS was co-transfected with gp91*phox* into HEK293 cells and analysed by western blot. **(F)** Normal or GFP-tagged c.327dup (p.Val110CysfsTer40) EROS DNA were cloned into a lentiviral backbone, transfected into HEK293T cells and transfection efficiency evaluated by flow cytometry. **(G)** The resulting lentivirus was used to transduce normal or GFP-tagged c.327dup (p.Val110CysfsTer40) EROS DNA into PLB985 EROS-/- cells, GFP expression was measured by flow cytometry and **(H)** expression of gp91*phox* and EROS was determined by western blot. Data are representative of n=3 experiments. N.S = non-specific band.

i. A frameshift from amino acid 110
ii. A premature stop codon at amino acid 140

Gnomad curates this variant as a putative loss-of-function variant ^6^. There are no homozygotes for this allele (or for any other EROS/*CYBC1* loss of function allele) in Gnomad, although heterozygotes are reported at a minor allele frequency of 0.00006545 in South Asian populations. The variant was not noted in any other ethnic group ^6^.

We performed experiments to examine whether this mutation was pathogenic. First, we used site directed mutagenesis to recapitulate the c.327dup (p.Val110CysfsTer40) mutation found in the patient in a pcDNA3.1 vector that expresses untagged EROS. We transfected the control and mutant vectors into HEK293 cells. In line with our previous studies, transfection with the control vector led to high levels of EROS expression ^4^. However, expression of the mutant vector did not lead to expression of any detectable EROS protein in multiple experiments and in a variety of transfection conditions (**Figure 2B**). The lack of expression of EROS protein by the mutant allele was not due to a failure of RNA expression or differing transfection efficiency; RNA expression of EROS at multiple exon junctions was equivalent following transfection of both control and mutant alleles (**Figure 2C**). We also saw no expression of the mutant allele compared with the wild-type allele when transfected into NIH3T3 cells, which express no endogenous EROS protein (**supplementary figure 1A)**. We reasoned that some mutant frameshifted EROS protein might still be made from the mutant allele but that it is no longer recognised by the anti-EROS antibody, which is specific for the C-terminus of the protein. To address this, we also engineered the mutant into a vector in which human EROS is fused with GFP at its C-terminus by site directed mutagenesis. Using GFP as a proxy for EROS expression we observed strong expression of the control vector but negligible expression of the mutant (**Figure 2D**). Similar results were also seen when we electroporated control and mutant GFP vectors into EROS deficient clone 14 cells (**supplementary figure 1B**).

Our previous work demonstrates the importance of EROS on gp91*phox* stability ^4^. Co-transfection of gp91*phox* with EROS leads to much greater expression of gp91*phox* than transfection of the gp91*phox* construct alone. Expression of the immature 58kDa gp91*phox* precursor was markedly increased by wild type EROS but not by the mutant vector (**Figure 2E)**.

A further way to assess whether a mutation in EROS leads to a functional effect is to use an EROS-GFP expressing lentivirus to reconstitute EROS expression in our previously described EROS-deficient PLB985 cell line, and determine whether this restores gp91*phox* expression (**supplementary figure 1B**). We created the c.327dup (p.Val110CysfsTer40) mutation in our lentiviral vector. As expected, this also led to negligible EROS protein expression and no reconstitution of gp91*phox* expression (**Figure 2F-H**).

## Discussion

CGD is a life threatening inborn error of immunity associated with severe morbidity and premature mortality. Early diagnosis is important as the prognosis is improved by factors such as appropriate prophylactic therapy, prompt treatment of severe fungal infections, recognition of inflammatory complications and management at specialist centres. Similarly, recent experience with haematopoietic stem cell therapy suggests that outcomes are excellent if patients are transplanted before they accrue severe complications in CGD, though more data are still required ^7,8^. Until recently, much of our knowledge of CGD was based on experience in high income countries such as the USA^9^, Western Europe^10^ and Japan^11^. Recently, there have been descriptions of cohorts in lower/middle income countries and these highlight distinct features of CGD such as the predominance of autosomal recessive CGD over X-linked CGD ^12,13^.

Mutations in EROS/*CYBC1* remain scarce and it is difficult to determine at present to what extent this form of IEI resembles classical CGD caused by mutations in gp91*phox*, p22*phox*, p47*phox* and p67*phox*. Notably, p40*phox* deficiency is a distinct clinical entity from those described above ^14,15^. Two major factors suggest that EROS/*CYBC1* deficiency may have distinct features:

i. EROS deficiency removes most but not all gp91*phox*-p22*phox*
ii. EROS controls the expression of other proteins such as P2×7 ^2,4,16^.

The patient we describe has a clinical history compatible with CGD including repeated episodes of granulomatous inflammation, severe and repeated infection with both bacterial and fungal pathogens and autoinflammatory phenomena. Septicaemia from *Salmonella* species is well recognised in CGD ^10,17,18^. Severe susceptibility to *Salmonella* Typhimurum infection in an *in vivo* screen was the means by which EROS was discovered ^2^. Osteomyelitis due to Cladophialophora has also been described in CGD ^19^. Mycobacterial infections are well recognised in CGD, particularly in areas of the world where they are endemic ^20^.

CGD is increasingly recognised in India. A previous report from a large centre in India showed that while they had described 17 cases before 2013, they described a further 21 between 2013 and 2017 ^21^. A recent multi-centre study of 236 patients in India expanded the phenotype further ^22^. This cohort focused largely on patients with mutations in *CYBB* (gp91*phox*) and *NCF1* (p47*phox*). In many respects, the patient that we describe here had a typical course for “classic” CGD both in terms of the localisation of infection and the organisms identified. For instance, she had pneumonia and lung inflammation, lymphadenitis, liver abscess and septicaemia. These are all well recognised features of CGD in general and also in the recent Indian cohort study. Similarly, the organisms identified in this patient are also well recognised CGD pathogens.

Our study is limited by the lack of availability of patient cells to confirm the absence of EROS, gp91*phox* and p22*phox* protein expression secondary to the mutation. This was caused by the logistical difficulties of transporting blood from the remote area in which the patient lived. Nevertheless, we were able to confirm the mutation by Sanger sequencing and demonstrate that it led to no detectable protein expression in several cell lines. These data, together with the typical clinical course, make it overwhelmingly likely that this is the third described mutation in EROS/*CYBC1* deficiency (CGD5).

Individuals that are heterozygous for the mutation we describe here have been observed in databases such as Gnomad, thus far restricted to people of South Asian ancestry. As such, we predict that there will be more cases of CGD5 caused by this mutation in EROS/*CYBC1*, both in India and elsewhere. The diagnosis should be considered in cases of CGD or other inborn errors of immunity, particularly when the generation of ROS is impaired without mutations in *CYBB, CYBA, NCF1, NCF2* and *NCF4*.

## Methods

### Consent

Informed consent for this study was obtained from the patient and family and ethical approval was granted by the Institutional Ethics Committee of Aster CMI Hospital, Bengaluru. IEC Regd. No. ECR/1084/Inst/KA/2018. The patient and their family are happy for the case to be published.

### Cell line culture

HEK293 and HEK293T cells were grown in complete DMEM (DMEM (Gibco) + 10% FBS (Pan biotech) + 1% penicillin-streptomycin-glutamine (ThermoFisher) + 1% sodium pyruvate (ThermoFisher). PLB985 and EROS-/- PLB985 cell lines were grown in complete RPMI (RPMI 1640 (Gibco), + 10% FBS + 1% penicillin-streptomycin-glutamine + 1% sodium pyruvate + 1% HEPES (ThermoFisher) +1% GlutaMAX (ThermoFisher)).

### DNA plasmids

DNA plasmids were obtained from Origene; untagged human EROS (SC324452), GFP tagged human EROS (RG200883) untagged gp91*phox* (SC122091), lentiEROS GFP tagged (RC200883L4) and equivalent empty vector (PS100093).

### Sanger sequencing

Patient and healthy donor DNA was isolated from whole blood using the Gentra PureGene kit (Qiagen), according to manufacturers’ instructions. DNA was subjected to standard PCR with the primers listed below. Amplified DNA was sequenced by GeneWiz and aligned using SnapGene.

Forward: AGTGCAGTGTAAAGATGG

Reverse: GTCCACAAACTCATCTCC

### Site-directed mutagenesis

A vector encoding EROS was generated using site-directed mutagenesis with primers listed below. Transfection was performed using QuikChange Lightning Multi Site-Directed Mutagenesis Kit (Agilent) as per the manufacturer’s instructions.

Forward (F327T): TGCTCCATGATGTCCGTGATTGTGAGCGTGGA

Reverse (R327A): TCCACGCTCACAATCACGGACATCATGGAGCA

### Transfection

HEK293/T cells were transfected with 1.2-1.8ng of DNA using lipofectamine RNAiMAX (Invitrogen) or lipofectamine2000 (Invitrogen). Cells were harvested at 48 or 72hrs for RNA, protein extraction and flow cytometry analysis.

### Nucleofection

EROS-/- PLB985 cells were nucleofected using the SF cell line 4D-Nucleofector X Kit L (Lonza), following manufacturer’s instructions. 1.2ng EROS-GFP or mutant EROS-GFP DNA was pulsed into cells using the EN-138 programme. Cells were harvested after 48hrs for flow cytometry analysis.

### Western blot

1×10^6^ cells were lysed with Pierce RIPA lysis buffer (ThermoFisher) containing 1x protein phosphatase inhibitor (ThermoFisher) and incubated for 30 minutes on ice. Debris was pelleted by centrifugation at 14000xg for 30 minutes at 4°C. Protein concentration was determined by BCA assay (ThermoFisher) according to manufacturer’s instructions. 20μg of protein was mixed with 4x LDS sample buffer (ThermoFisher) + 10% reducing agent (Invitrogen) and denatured at 72°C for 10 minutes. Samples were run on NuPAGE 4-12% Bis-Tris gels (ThermoFisher) in 1x MOPS running buffer (Formedium) at 100-150v for 60-90mins. Proteins were transferred to a nitrocellulose membrane (ThermoFisher) in transfer buffer (3.05g Tris base (Sigma) + 14.45g Glycine (Sigma) + 800ml dH2O + 200ml methanol) and ran for 1 hour 15 mins at 100v. The membrane was blocked in 5% skim milk (Marvel) in TBST for 1 hour at room temperature. Membrane was cut and probed with appropriate primary antibody; actin (1:2500, abcam), EROS (1:1000, Atlas), gp91*phox* (1:1000, Santa Cruz), vinculin (1:1000, CST) in blocking buffer overnight at 4°C on a roller. Membranes were washed 3x in TBST for 10 minutes and incubated with appropriate secondary antibody (anti-rabbit IgG horseradish peroxidase, 1:10000 (CST) or anti-mouse IgG horseradish peroxidase, 1:10000 (CST)) in blocking buffer for 1 hour at room temperature. The blots were developed with SuperSignal West Pico PLUS (ThermoFisher) or SuperSignal West Femto PLUS (ThermoFisher) according to manufacturer’s instructions and imaged on a ChemiDoc (BioRad).

### qPCR

1×10^6^ cells were resuspended in RLT buffer (QIAGEN) + 10% β-mercaptoethanol (Sigma) on ice. RNA was extracted using QIAGEN RNAeasy mini kit according to manufacturer’s instructions. cDNA synthesis was carried out using superscript IV VILO master mix (Invitrogen) according to manufacturer’s instructions using 1μg of RNA. qPCR performed using TaqMan Fast advanced master mix (ThermoFisher) according to manufacturer’s instructions. EROS primers used were Hs00978805 exon 2-3, Hs00978806 exon 3-4, Hs00978808 exon 5-6 and housekeeping HPRT Hs02800695 (ThermoFisher).

### Flow cytometry

GFP tagged samples were acquired on a LSRFortessa (BD).

### Generation of lentivirus

Lentiviral mix was added to HEK293T cells at a ratio of 4 lentiviral plasmid: 3 psPAX2 (Addgene): 1 pMD2.G (Addgene), using 7μg of lentiviral plasmid in lipofectamine2000. Cells were incubated at 37°C, 5% CO_2_ for 48hrs. The viral supernatant was harvested and filtered and concentrated using LentiX concentrator (Takara). Transfected HEK293T cells were harvested and analysed by flow cytometry to assess transfection efficiency.

### Lentiviral transduction

1×10^5^ PLB985 and EROS-/- PLB985 cells were transduced with viral supernatant plus 8μg/ml polybrene (Merck) and incubated at 37°C, 5% CO2 for 72 hrs. Transduction efficiency was analysed by flow cytometry to determine GFP expression. Remaining cells were expanded in culture and harvested for western blot.

Figures were created using Biorender.

## Data Availability

All data produced in the present work are contained in the manuscript

## Abbreviations

ATT: Anti-Tuberculosis Treatment
CGD: Chronic Granulomatous Disease
DHR: Dihydrorhodamine
EROS: Essential for Reactive Oxygen Species
IEI: Inherited Error of Immunity
NADPH: Nicotinamide adenine dinucleotide phosphate
ROS: Reactive Oxygen Species

## Funding Statement

This work was supported by funding from a Wellcome-Beit Prize Trust Clinical Research Career Development Fellowship (20661206617/A/17/Z and 206617/A/17/A) and the Sidharth Burman endowment. MCP is a Wellcome Trust Senior Fellow in Clinical Science (212252/Z/18/Z) and THM is supported by this fellowship.

We acknowledge support from the National Institute for Health Research (NIHR) Biomedical Research Centre based at Imperial College Healthcare National Health Service Trust and Imperial College London, and from the NIHR Clinical Research Network. The views expressed are those of the authors and not necessarily those of the National Health Service, the NIH, or the Department of Health.

We thank the LMS/NIHR Imperial Biomedical Research Centre Flow Cytometry Facility for their support.

---

**Supplemental Figure 1:**
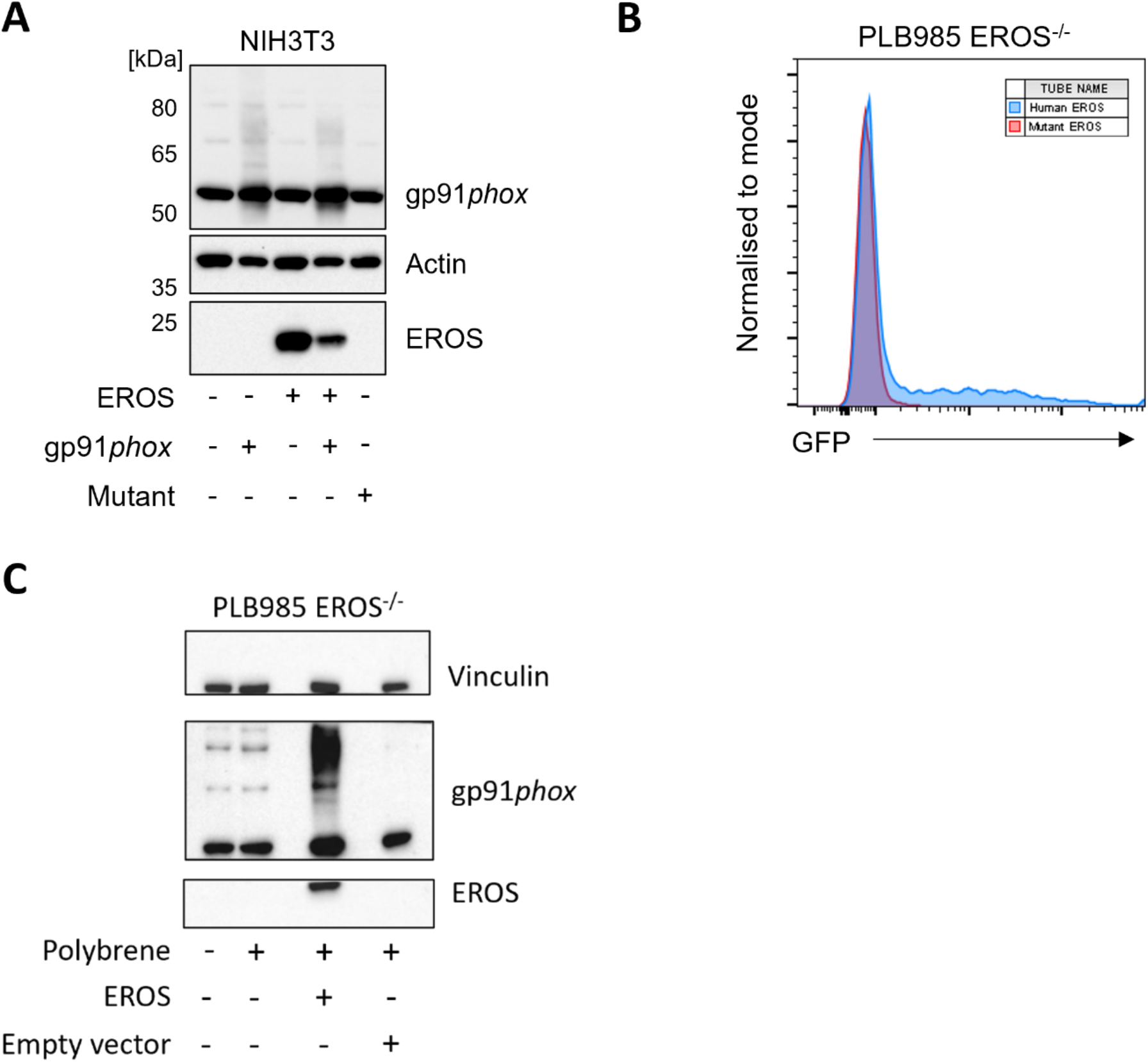
**(A)** Normal and c.327dup (p.Val110CysfsTer40) EROS DNA was transfected into NIH3T3 cells and EROS and gp91*phox* protein expression was measured by western blot **(B)** GFP-tagged c.327dup (p.Val110CysfsTer40) and normal EROS DNA were nucleofected into PLB985 EROS-/- cells. **(C)** Transduction of normal EROS successfully reconstituted gp91*phox* expression in PLB985 EROS-/- cells. Actin or vinculin used as loading control. Data are representative of n=3 experiments.

## Acknowledgements

We thank the patient and family for their contributions to this research. We thank Dr Kerry Rostron for facilitating several logistic aspects of this study.

## Declarations

None of the authors has any potential financial conflict of interest related to this manuscript. Declarations of interest: none - manuscript file

